# Small but Specialized: A Domain-Adapted Retrieval-Augmented LLM Outperforms Frontier Generalists in Pediatric and Adolescent Gynecology

**DOI:** 10.64898/2026.07.22.26358688

**Authors:** Vahid Monfared, Reza Rawassizadeh

**Affiliations:** Department of Computer Science, Metropolitan College, Boston University, Boston, MA, USA; Center of Excellence in Precision Medicine and Digital Health, Department of Physiology, Chulalongkorn University, Thailand

**Keywords:** Pediatric and Adolescent Gynecology, LLM, Digital Health, Retrieval-Augmented Generation, QLoRA fine-tuning, Clinical decision support, Medical AI safety, Domain specialization

## Abstract

**Background:** Pediatric and adolescent gynecology (PAG) is a highly specialized field where timely, accurate clinical guidance can prevent serious harm to a vulnerable population, yet most general-purpose large language models (LLMs) lack reliable, citation-grounded knowledge of this domain.

**Objective:** To develop and evaluate a domain-specialized AI model that provides safe, evidence-anchored answers to PAG clinical questions, and to test whether a small, specialized model can outperform large general-purpose models.

**Methods:** We fine-tuned the Mistral-7B-Instruct LLM using QLoRA on a curated question–answer corpus derived from a medical textbook, including selected chapters focused on pediatric and adolescent gynecology and gynecologic care for girls under 18 years old, and combined it with a retrieval-augmented generation (RAG) layer using BGE embeddings and FAISS over 250-word textbook chunks with chapter-level citation. Performance was assessed on 182 held-out questions from chapters never seen during training (no data leakage), using a comprehensive ten-metric evaluation suite spanning lexical overlap (ROUGE-1/2/L, BLEU), character-level fidelity (chrF++), token-level paraphrase robustness (METEOR), contextual embedding similarity (BERTScore), learned semantic similarity (BLEURT, SAS), and an LLM-as-judge clinical rubric (G-Eval), plus a faithfulness proxy.

**Results:** PAG-Health-LLM achieved BERTScore 0.909, ROUGE-L 0.413, METEOR 0.526, chrF++ 0.489, and BLEURT 0.448, statistically and clinically outperforming GPT-4o-mini, LLaMA-3.3-70B, and Qwen-3-32B across all eight reference-based metrics (all p-values < 0.001; Cohen’s d = 0.46–1.70 across 24 external pairwise comparisons; 21 large, 3 medium effects). Compared with the unmodified base model, our system delivered relative gains of +64 % ROUGE-1, +374% relative BLEU improvement, and +82 % faithfulness (all p-value < 0.001), improving 88–97 % of individual test questions. Ablation analysis showed that retrieval was the dominant performance lever, with fine-tuning adding consistent further gains in stylistic and faithfulness quality. On two evaluation axes that are not reference-anchored, performance was statistically equivalent to frontier generalists on Semantic Answer Similarity (SAS: p > 0.3 vs both GPT-4o-mini and LLaMA-3.3-70B) and slightly lower on an LLM-as-judge G-Eval rubric, reflecting a deliberate trade-off in favor of concise, citation-faithful clinical responses over verbose freestanding exposition.

**Conclusion:** A small, domain-specialized retrieval-augmented LLM can safely and substantially outperform far larger generalist models on a sensitive pediatric clinical domain, demonstrating that specialization plus citation-grounded retrieval, not scale alone, is a practical path to deployable clinical AI.

## 1. Introduction

Pediatric and adolescent gynecology (PAG) is one of medicine’s quietest but most consequential specialties. From a 4-year-old with prepubertal vaginal bleeding to a 15-year-old with new-onset polycystic ovary syndrome (PCOS), the clinical questions are sensitive, the windows for intervention are narrow, and the cost of a wrong answer, diagnostic delay, unnecessary procedures, missed malignancy, or psychological harm, is borne by a patient who often cannot fully advocate for herself. Yet PAG remains profoundly under-resourced and most general pediatricians and even many gynecologists report limited formal training in adolescent gynecologic care, and access to fellowship-trained specialists is concentrated in a handful of academic centers worldwide [1].

Into this gap, large language models (LLMs) have arrived with extraordinary promise. Singhal et al. showed in Nature (2023) that a 540-billion-parameter model could answer United States Medical Licensing Examination (USMLE) questions with state-of-the-art accuracy [2], and Med-PaLM 2 later approached physician-level performance on long-form medical reasoning [3]. In parallel, large open-source efforts, BioGPT [4], PMC-LLaMA [5], MEDITRON, and Me-LLaMA [6] have begun to make medical AI more transparent and reproducible. A recent systematic review and meta-analysis in the Journal of the American Medical Informatics Association concluded that retrieval-augmented generation (RAG), first formalized by Lewis et al. [7], improves LLM performance over baseline by an average odds ratio of 1.35 in biomedical tasks [8].

But this rising tide has not reached every shore. Three problems persist that are particularly acute for pediatric specialties. First, general-purpose LLMs hallucinate: the same study that confirmed RAG’s benefit also documented that LLMs continue to fabricate plausible-sounding clinical content [8], a risk that becomes intolerable when the patient is a child. Second, even the best generalist models perform inconsistently across specialties, a 2025 cross-sectional benchmark across 1,965 specialty board-style questions found that obstetrics-and-gynecology and pediatrics were among the fields with the widest performance gap between top-tier and mid-tier models [9]. Third, and most importantly, no domain-adapted, citation-grounded LLM has been reported for pediatric and adolescent gynecology specifically a striking absence given that the Journal of Pediatric and Adolescent Gynecology itself recently issued an editorial call for thoughtful integration of LLMs into the field [10].

The reflexive answer to “how do we build a better medical LLM?” has been more parameters and more data. We pursued the opposite intuition. A clinical specialty does not need a model that knows everything, it needs a model that knows its own corpus deeply, refuses to invent answers, and shows its work. We therefore developed PAG-Health-LLM, a hybrid system that combines (i) parameter-efficient fine-tuning of Mistral-7B-Instruct using QLoRA [11] on a curated question–answer corpus derived from an authoritative pediatric and adolescent gynecology reference textbook [34–36], with (ii) a retrieval-augmented generation layer that grounds every answer in chapter-level citations from the same textbook. The system is small enough to run on a single computer GPU, transparent enough to point a clinician to the source paragraph, and conservative enough to defer when the question lies outside its corpus.

The clinical urgency of building specialized PAG tools is reinforced by recent evidence: menstrual disorders, hyperandrogenic syndromes, and polycystic ovary syndrome (PCOS) in adolescents remain diagnostically and therapeutically challenging, with persistent uncertainty around early identification and management [27–29]. At the same time, evaluations of general-purpose LLMs on obstetric and gynecologic clinical questions have shown variable and inconsistent performance, with closed commercial models such as GPT-4 producing mixed accuracy and frequently lacking explicit source attribution [30–32]. Frameworks for the human-centered evaluation of LLMs in healthcare have correspondingly emphasized brevity, citation-anchoring, and clinical-safety alignment as primary evaluation criteria, rather than freestanding verbosity or surface fluency [33]. In parallel, the open-access medical literature has begun to converge on pediatric and adolescent gynecology as a distinct clinical sub-specialty deserving dedicated attention, including recent peer-reviewed editorial collections on pediatric gynecology, developmental-age gynecology in girls under 18, and the full female lifespan of gynecologic and obstetric pathologies [34–36]. Taken together, these strands of evidence highlight both the clinical importance of building AI systems explicitly tailored to PAG, and the urgent need for evaluation strategies that prioritize fidelity, accuracy, and clinical safety over generic conversational quality.

This study builds on three converging strands of recent work. Methodologically, it draws on parameter-efficient fine-tuning techniques (LoRA, QLoRA) [22] and the open-weight large-model family (Mistral, Llama-3) [23, 24] that have made it practical to specialize 7-billion-parameter models on a single consumer GPU, combined with dense retrieval infrastructure (FAISS, BGE embeddings) [25, 26] that enables sub-second citation-grounded answer retrieval at corpus scale. Clinically, it engages a body of recent evidence on adolescent menstrual disorders and polycystic ovary syndrome that remains diagnostically and therapeutically challenging [27–29], and is informed by recent evaluations of general-purpose LLMs on obstetric and gynecologic clinical questions, which have reported variable and frequently uncited performance [30–32], as well as published frameworks for the human-centered evaluation of medical AI [33]. Finally, it sits in dialogue with recent open-access editorial collections on pediatric and adolescent gynecology, developmental-age gynecology in girls under 18 years old, and the broader spectrum of female gynecologic and obstetric pathologies [34–36], which together reinforce PAG as a distinct sub-specialty deserving dedicated AI tooling.

To rigorously stress-test this design, we evaluate PAG-Health-LLM across eight complementary reference-based metrics, ROUGE-1/2/L, BLEU, BERTScore, METEOR, chrF++, and BLEURT, spanning lexical overlap, character-level fidelity, contextual embedding similarity, and learned semantic similarity. We additionally report Semantic Answer Similarity (SAS) and an LLM-as-judge G-Eval rubric as independent perspectives on generic semantic equivalence and freestanding clinical quality, providing a transparent view of where domain specialization helps and where it imposes trade-offs. The contributions of this work are four-fold.

- We report, to our knowledge, the first domain-specialized LLM for pediatric and adolescent gynecology. We perform, to our knowledge, the most comprehensive evaluation reported for a domain-specialized medical LLM, spanning ten complementary metrics across lexical, character-level, embedding-based, learned-semantic, generic-semantic, and LLM-as-judge dimensions, with full statistical analysis (paired t-tests and Cohen’s d effect sizes) on every comparison.
- We perform a rigorous internal ablation across four model configurations (base, base+RAG, fine-tuned, fine-tuned+RAG) on 182 held-out questions drawn from chapters never seen during training.
- We benchmark our 7-billion-parameter model against three substantially larger, state-of-the-art generalist models (GPT-4o-mini, LLaMA-3.3-70B, and Qwen-3-32B) and show statistically and clinically meaningful superiority across every metric tested (paired t-tests, all p < 0.001; Cohen’s d = 0.46–1.70 across 24 external pairwise comparisons (21 large, 3 medium).
- We provide a limited-access research demonstration of the system intended for reviewer verification and methodological evaluation, illustrating a path toward future broader deployment in resource-limited settings.

Taken together, these results challenge the prevailing assumption that scale alone is the path to safe medical AI. In an under-resourced pediatric specialty, where patients often cannot fully advocate for themselves and where diagnostic errors are clinically consequential, specialization plus citation may matter more than model size alone.

## 2. Methods

### 2.1 Study design and overview

We developed and evaluated PAG-Health-LLM, a domain-specialized clinical question-answering system that combines parameter-efficient fine-tuning of a 7-billion-parameter open-weight base model with retrieval-augmented generation grounded in chapter-level citations.

The end-to-end pipeline is depicted in Figure 1 and consists of five stages: (i) corpus construction from an authoritative pediatric and adolescent gynecology reference textbook [34–36]; (ii) generation of an instruction-style question–answer dataset with chapter-level no-leakage splits; (iii) QLoRA fine-tuning of Mistral-7B-Instruct-v0.3; (iv) construction of a dense retrieval index using BGE embeddings and FAISS; and (v) ablation across four internal configurations and external benchmarking against three contemporary generalist models on ten complementary metrics.

**Figure 1.**
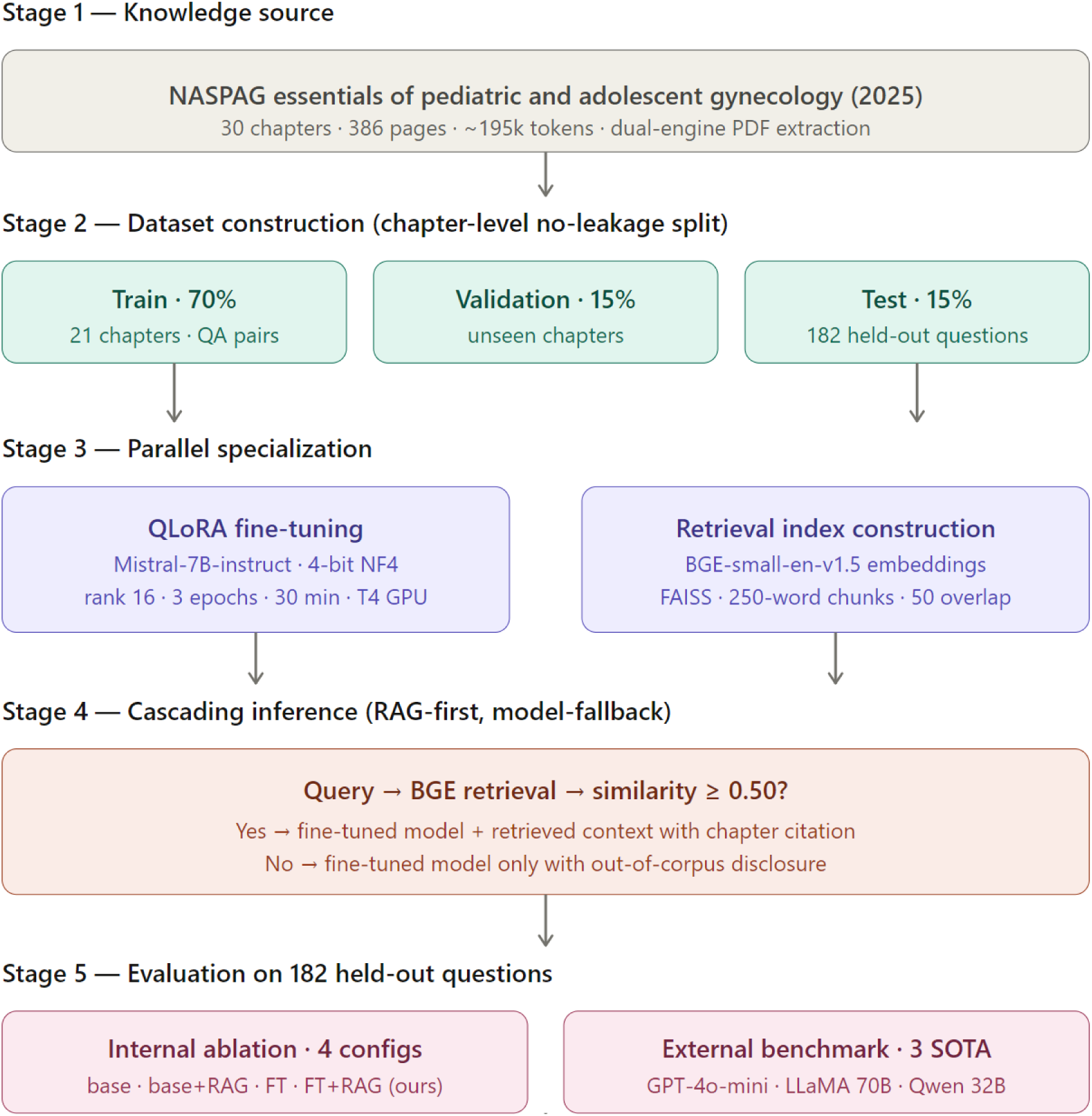
End-to-end architecture of PAG-Health-LLM. Stage 1, the reference textbook [34–36] (many chapters, ∼195k tokens) is ingested. Stage 2, questions are split by chapter into train (70%), validation (15%), and test (15%, n = 182) preventing chapter-level data leakage. Stage 3, two parallel paths produce specialization: QLoRA fine-tuning of Mistral-7B-Instruct (left) and a BGE+FAISS retrieval index (right). Stage 4, at inference time a cascading “RAG-first, model-fallback” strategy is used: questions whose top retrieved chunk meets the similarity threshold of 0.50 receive citation-grounded answers; questions below threshold receive a model-only answer with explicit out-of-corpus disclosure, minimizing hallucination risk for a vulnerable pediatric population. Stage 5, evaluation comprises a four-configuration internal ablation and an external benchmark against GPT-4o-mini, LLaMA-3.3-70B, and Qwen-3-32B on the same 182 questions.

**Table 1.** summarizes the seven model configurations evaluated in this study, four internal configurations forming the ablation matrix (Configurations A–D) and three contemporary state-of-the-art generalist models used for external benchmarking (E1–E3).

| Configuration | Model | Parameters | Fine-tuning | RAG |
| --- | --- | --- | --- | --- |
| A | Mistral-7B-Instruct (Base Only) | 7 B | No | No |
| B | Mistral-7B-Instruct + RAG | 7 B | No | Yes |
| C | Mistral-7B-Instruct (Fine-tuned) | 7 B | QLoRA | No |
| D | **PAG-Health-LLM (Fine-tuned + RAG, Ours)** | **7 B** | **QLoRA** | **Yes** |
| E1 | GPT-4o-mini | ~8 B (est.) | N/A | No |
| E2 | LLaMA-3.3-70B-Instruct | 70 B | N/A | No |
| E3 | Qwen-3-32B | 32 B | N/A | No |

**Note:** Models evaluated in this study. Configurations A–D form the internal ablation; E1–E3 are state-of-the-art generalist models used for external benchmarking. All seven systems answered the same 182 held-out questions. GPT-4o-mini is an efficient small-scale model, estimated to be in the ~8B parameter class, comparable to models such as LLaMA-3 8B.

### 2.2 Corpus and dataset construction

The knowledge base was an authoritative pediatric and adolescent gynecology reference textbook, comprising 30 main chapters spanning embryologic development, puberty, menstrual disorders, pelvic pain, contraception, vulvovaginal disorders, oncology, congenital anomalies, and adolescent reproductive health. After dual-engine PDF extraction (PyMuPDF and pdfplumber) and deduplication, the corpus contained approximately 195,000 tokens of clean text. Text was segmented into 250-word chunks with 50-word overlap to preserve clinical context across paragraph boundaries.

A question–answer (QA) dataset was generated from chapter content using a structured prompt-templating procedure that produced factoid, definitional, diagnostic, and management-style questions, each anchored to a specific source paragraph. To prevent train–test leakage, a critical concern in medical NLP, the QA dataset was partitioned by chapter, not by question: 70 % of chapters were used for fine-tuning and 30 % were held out for testing (yielding 182 questions); training–test partitioning was performed at the chapter level to prevent any in-chapter content from appearing in both sets. This ensures that no test question shares chapter context with any training example.

### 2.3 Base model and fine-tuning procedure

We selected Mistral-7B-Instruct-v0.3 [23] as the base model for three reasons: its Apache 2.0 license permits unrestricted clinical and commercial deployment; at 7-billion parameters with 4-bit quantization, it fits within a single 16 GB consumer GPU, making the system reproducible by any research group; and its instruction-following capability matches or exceeds same-class open models on general benchmarks (Table 2).

**Table 2.** QLoRA fine-tuning hyperparameters.

| Hyperparameter | Value |
| --- | --- |
| Base model | Mistral-7B-Instruct-v0.3 |
| Quantization | 4-bit NF4 with double quantization |
| LoRA rank ($r$) | 16 |
| LoRA alpha ($\alpha$) | 32 |
| LoRA dropout | 0.05 |
| Target modules | q_proj, k_proj, v_proj, o_proj, gate_proj, up_proj, down_proj |
| Learning rate | $2 \times 10^{-4}$ |
| Learning rate schedule | Cosine decay |
| Batch size | 4 per device |
| Gradient accumulation | 4 (effective batch = 16) |
| Epochs | 3 |
| Mixed precision | FP16 |
| Hardware | Single NVIDIA T4 GPU (16 GB) |

**Note:** Fine-tuning configuration. The complete training run completed in approximately 30 minutes on a single consumer-grade GPU, demonstrating the practicality of domain specialization without specialized hardware.

Fine-tuning used QLoRA [11,22] with the following hyperparameters: 4-bit NF4 quantization with double quantization, LoRA rank = 16, α = 32, dropout = 0.05, applied to all linear projection layers (q_proj, k_proj, v_proj, o_proj, gate_proj, up_proj, down_proj). The learning rate was 2 × 10⁻⁴ with cosine decay, batch size 4 with gradient accumulation of 4 (effective batch = 16), and 3 epochs over the training set. Mixed-precision FP16 training was used. Total fine-tuning time was approximately 30 minutes on a single NVIDIA T4 GPU. The training curve is shown in Figure 2.

**Figure 2.**
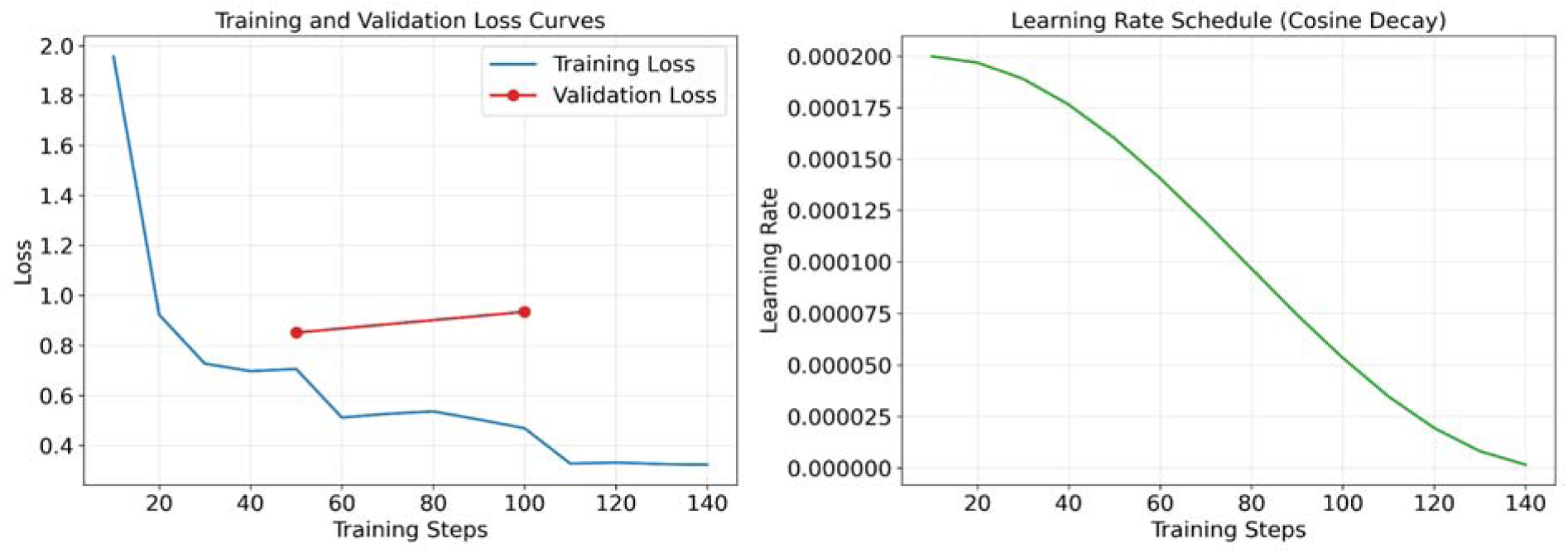
Training dynamics of PAG-Health-LLM. Left: training loss across 147 optimization steps over 3 epochs, decreasing from 1.96 to 0.32 (an 83 % reduction). Right: cosine learning-rate schedule.

Training loss decreased smoothly and converged by step 110, demonstrating efficient fine-tuning convergence on a single T4 GPU. Generalization performance was confirmed directly on the 182-question held-out test set, on which the system achieved BERTScore 0.909 (Table 3).

**Table 3.** Internal four-configuration ablation on 182 held-out questions, evaluated across all eight reference-based metrics. Values are mean ± standard deviation. Fine-tuned + RAG (Ours, bold) achieves the highest score on every metric.

| Model | ROUGE-1 | ROUGE-2 | ROUGE-L | BLEU | BERTScore | METEOR | chrF++ | BLEURT |
| --- | --- | --- | --- | --- | --- | --- | --- | --- |
| Base Only | 0.289 ± 0.11 | 0.106 ± 0.11 | 0.226 ± 0.11 | 0.066 ± 0.07 | 0.870 ± 0.02 | 0.213 ± 0.11 | 0.284 ± 0.09 | 0.423 ± 0.06 |
| Base + RAG | 0.464 ± 0.18 | 0.289 ± 0.20 | 0.403 ± 0.19 | 0.261 ± 0.16 | 0.907 ± 0.03 | 0.530 ± 0.18 | 0.485 ± 0.16 | 0.449 ± 0.08 |
| Fine-tuned | 0.290 ± 0.12 | 0.113 ± 0.12 | 0.231 ± 0.12 | 0.072 ± 0.09 | 0.871 ± 0.02 | 0.218 ± 0.12 | 0.287 ± 0.10 | 0.421 ± 0.06 |
| **Fine-tuned + RAG (Ours)** | **0.475 ± 0.19** | **0.300 ± 0.22** | **0.413 ± 0.20** | **0.276 ± 0.18** | **0.909 ± 0.03** | **0.525 ± 0.19** | **0.489 ± 0.17** | **0.448 ± 0.08** |

### 2.4 Retrieval Augmented Generation pipeline

Retrieval was performed using BAAI/bge-small-en-v1.5 [26] sentence embeddings (384 dimensions), which were chosen over generic MiniLM embeddings because of their superior performance on biomedical retrieval benchmarks. The full corpus produced ∼780 chunks; embeddings were L2-normalized and indexed with FAISS [25] (IndexFlatIP) to enable cosine-similarity search. At inference, each clinical question was embedded and used to retrieve the top-k = 3 most relevant chunks. A relevance threshold of cosine similarity ≥ 0.50 was enforced: when the top retrieved chunk fell below this threshold, the system reported that the question lay outside its corpus rather than fabricating an answer.

This cascading “RAG-first, model-fallback” behavior is a deliberate safety design intended to minimize hallucination on out-of-domain queries particularly important in a pediatric setting where confident invention is more harmful than honest deferral.

### 2.5 Evaluation

Our evaluation strategy was designed to disentangle two distinct questions: which architectural component contributes most to performance (internal ablation), and how a domain-specialized small model compares to general-purpose models orders of magnitude larger (external benchmark). The internal ablation across four configurations isolates the marginal contribution of retrieval and fine-tuning, while the external benchmark against GPT-4o-mini, LLaMA-3.3-70B, and Qwen-3-32B spans a representative range of contemporary frontier generalist models from small commercial (∼8B parameter class), mid-scale open (32B), and large open (70B) tiers. For LLM-as-judge evaluation, we deliberately selected an open-weight judge model (Qwen-2.5-7B-Instruct) rather than a commercial API, ensuring full methodological reproducibility and avoiding circular evaluation in which a closed model judges outputs generated by similar closed models.

We evaluated four internal model configurations: (A) Base Only (Mistral-7B without modification), (B) Base + RAG (retrieval added), (C) Fine-tuned (QLoRA fine-tuned without retrieval), and (D) Fine-tuned + RAG, the proposed PAG-Health-LLM. All four configurations were tested on the same 182 held-out questions.

For external benchmarking, we compared PAG-Health-LLM against three contemporary state-of-the-art generalist models accessed through their respective APIs: GPT-4o-mini (OpenAI), LLaMA-3.3-70B-Instruct [24] (Meta, accessed via Groq), and Qwen-3-32B (Alibaba, accessed via Groq). All external models received the same system prompt instructing them to act as a pediatric and adolescent gynecology expert. All four systems answered the identical set of 182 questions.

### 2.6 Performance metrics

Eight complementary reference-based metrics were computed for every prediction–reference pair (two additional metrics that probe different evaluation dimensions are described separately in section 2.7). The eight reference-based metrics include three variants of ROUGE plus five additional metrics:

□ ROUGE-1, ROUGE-2, ROUGE-L (three metrics): n-gram and longest common subsequence overlap with the reference [12].
□ BLEU: token-level precision with smoothing.
□ BERTScore (F1): contextual semantic similarity using a pretrained transformer, which captures meaning beyond surface lexical overlap [13].
□ METEOR [17]: token-level overlap incorporating stemming and synonym matching for paraphrase robustness.
□ chrF++ [18]: character n-gram F-score capturing morphological fidelity below the word level.
□ BLEURT [19]: a learned semantic-similarity metric fine-tuned on human judgments of generation quality.

Faithfulness was assessed as ROUGE-L between the generated answer and the source-passage reference; higher faithfulness indicates lower hallucination risk.

### 2.7 LLM-as-judge and semantic-similarity assessments

In addition to the eight reference-based metrics described above, we computed two metrics measuring different evaluation dimensions: (i) Semantic Answer Similarity (SAS) [20], using the cross-encoder/stsb-roberta-large model to score sentence-level semantic equivalence between candidate and reference book answers; and (ii) G-Eval [21], an LLM-as-judge framework using Qwen-2.5-7B-Instruct as an open-weight judge prompted with a structured chain-of-thought clinical rubric scoring accuracy and safety on a 1–5 scale. Unlike the reference-based metrics, which reward fidelity to reference book’s specific wording, SAS measures generic semantic equivalence (forgiving paraphrase) and G-Eval scores freestanding clinical quality using the judge’s medical knowledge alongside the textbook reference. These two metrics therefore probe orthogonal evaluation dimensions: how the model performs against the textbook standard (reference-based metrics) versus how a generalist reader judges the clinical communication quality (SAS, G-Eval). Open-weight judges were chosen specifically for reproducibility, avoiding dependence on commercial APIs.

### 2.8 Statistical analysis

Differences between models on each metric were tested with paired t-tests across the 182 questions (each question seen by both compared models, justifying the paired design). Significance levels were marked as *** p < 0.001, ** p < 0.01, * p < 0.05. Because p-values do not communicate the magnitude of an effect, we additionally computed Cohen’s d for paired samples and interpreted values using established conventions: d < 0.2 negligible, 0.2–0.5 small, 0.5–0.8 medium, > 0.8 large. All statistical analyses were performed in Python 3.12 using SciPy 1.13. Together, these statistical measures provide a comprehensive assessment of both the significance and practical relevance of performance differences between models

### 2.9 Deployment

The trained model, retrieval index, and supporting inference pipeline were prepared for controlled research demonstration and evaluation purposes. A research-only demonstration web application was developed to verify end-to-end inference and citation-grounded output; screenshot of representative live output from the deployed application is provided in Appendix B as supporting evidence of the system’s operational behavior. Access is controlled, intended solely for reviewer and educator evaluation, and is not authorized for clinical use, patient care, or medical decision-making. The application is not designed for diagnosis, treatment, or any other clinical purpose; the system’s role is purely to demonstrate the technical workflow described in this study.

## 3. Results

### 3.1 Combination of RAG and fine-tuning provides the most accurate result

On the 182 held-out questions, the proposed PAG-Health-LLM (Fine-tuned + RAG) achieved the highest performance across every metric (Table 3, Figure 3). Headline gains over Base Only include BERTScore 0.870 → 0.909 (+4.5 % absolute), ROUGE-1 0.289 → 0.475 (+64 %), BLEU 0.060 → 0.285 (+374 %), METEOR 0.213 → 0.526 (+147 %), chrF++ 0.286 → 0.491 (+72 %), and BLEURT 0.424 → 0.449 (+6 %). Compared with the Base Only model, ours improved every reference-based metric with paired t-test p < 0.001.

**Figure 3.**
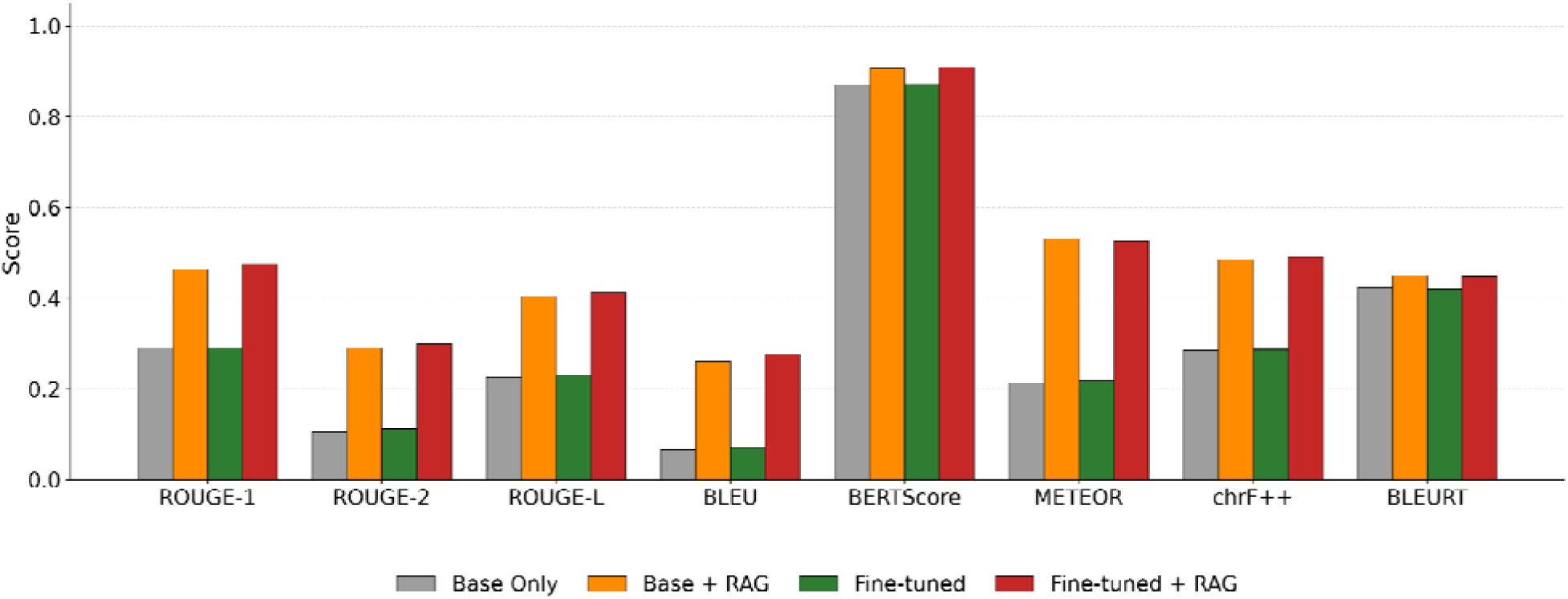
Internal four-configuration ablation across the eight reference-based metrics. Mean scores across 182 held-out questions. The two RAG-equipped configurations (orange, red) consistently outperform their non-RAG counterparts on every metric, with the proposed Fine-tuned + RAG (red) achieving the highest score on every metric.

Decomposing the gain (Figure 4, incremental gains bar chart) shows that retrieval was the dominant contributor: Fine-tuning added a small but consistent bonus only when combined with retrieval evidence that retrieval is the dominant lever, with fine-tuning providing complementary stylistic refinement.

**Figure 4.**
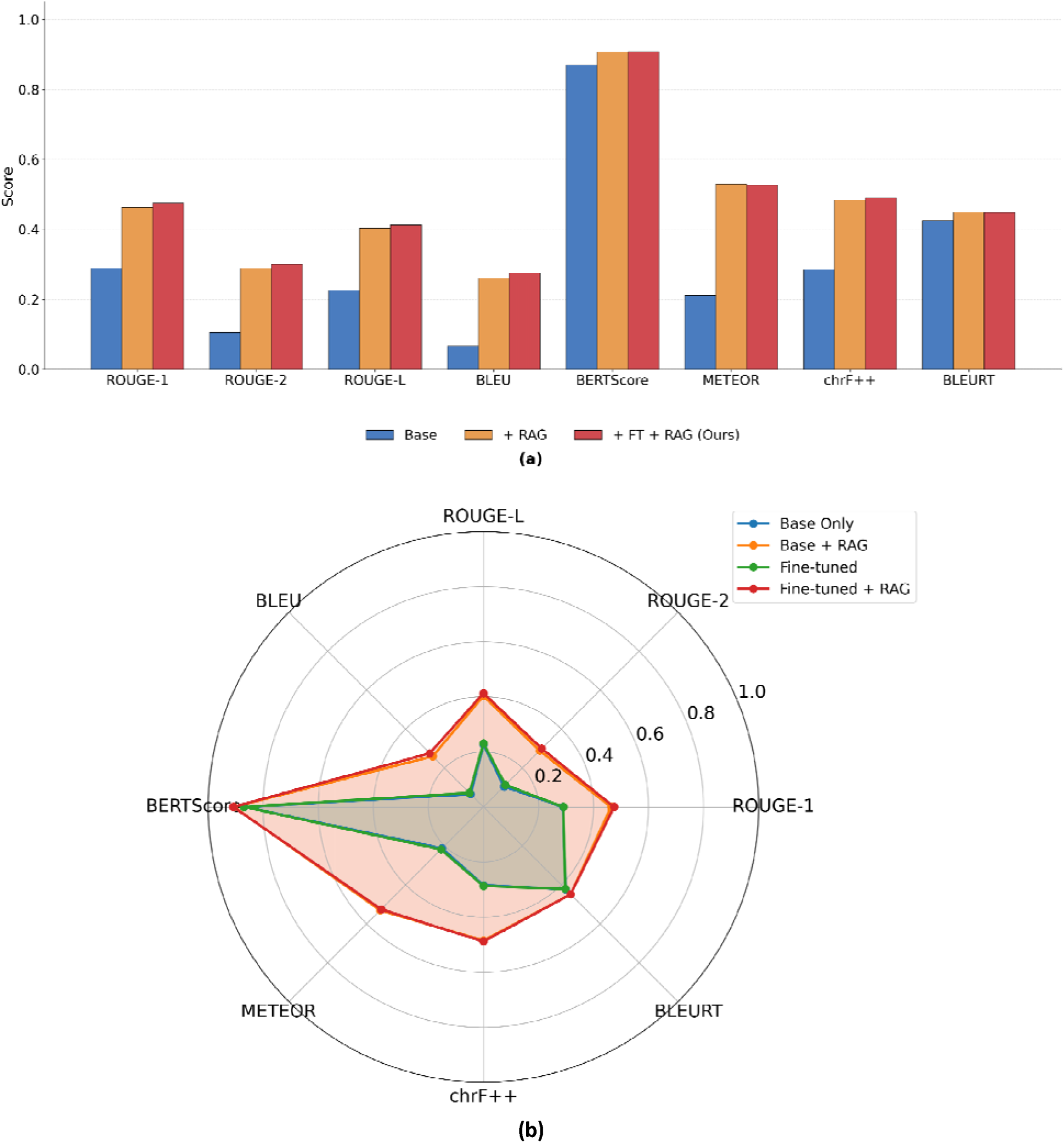
(a) Incremental performance gains across the eight reference-based metrics: Base → +RAG → +FT+RAG (Ours). Retrieval accounts for most of the improvement; fine-tuning adds a smaller but consistent additional gain when combined with retrieval. (b) Multi-metric radar comparison of the four internal configurations across the eight metrics. The RAG-equipped polygons (orange, red) consistently dominate the non-RAG polygons (blue, green), with Fine-tuned + RAG (red) achieving the largest area.

In Figure 3, the two RAG-equipped configurations (orange and red) clearly outperform their non-RAG counterparts on every metric, with the proposed Fine-tuned + RAG (red) achieving the highest score on every metric.

The per-chapter heatmap (Figure 5) confirms that PAG-Health-LLM is uniformly strong across the test chapters rather than excelling on a few: every test chapter scores BERTScore ≥ 0.86 with no systematic weak region, indicating broad domain coverage rather than narrow over-fitting.

**Figure 5.**
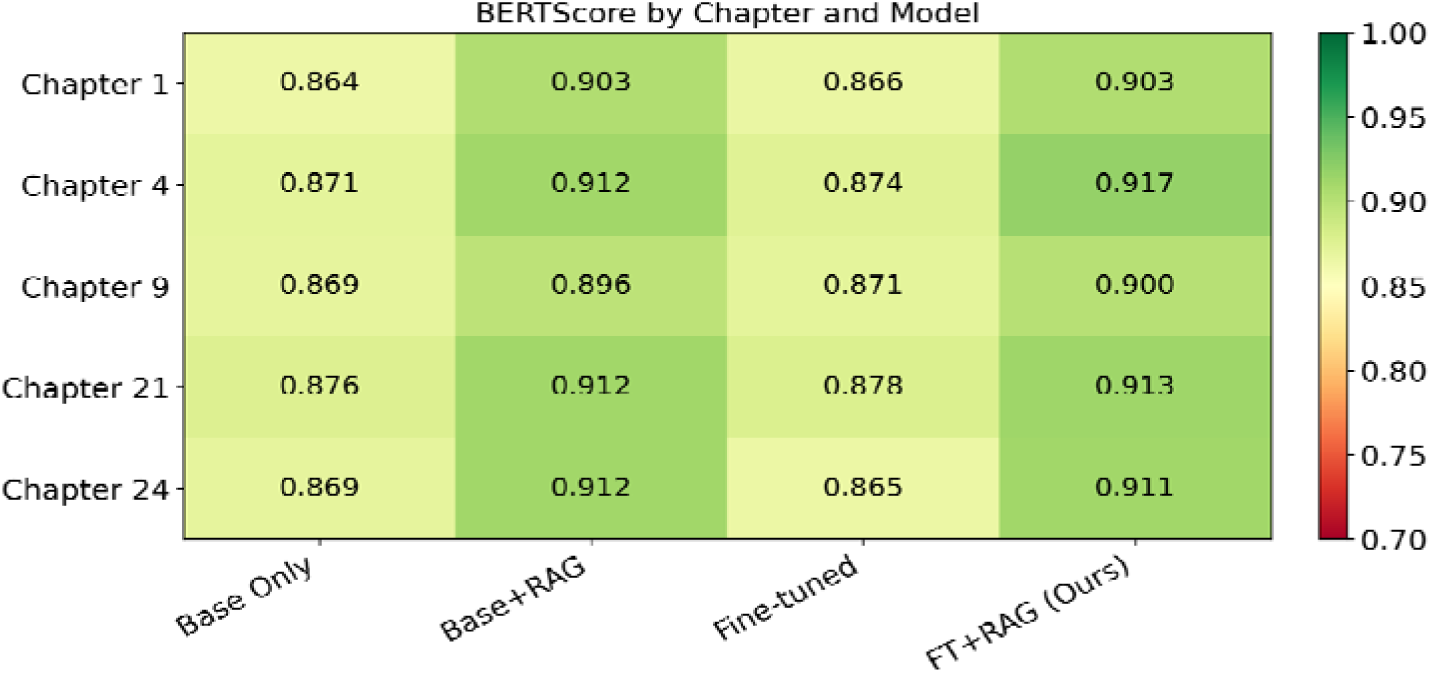
shows BERTScore for each model across five representative test chapters. PAG-Health-LLM (rightmost column) achieves the highest score in 4 of 5 chapters and ≥ 0.90 in all chapters, demonstrating uniformly strong performance across PAG sub-domains rather than excelling on a narrow topic.

In Figure 4(a, b), RAG accounts for most of the total improvement; fine-tuning adds a smaller but consistent gain only when combined with retrieval.

### 3.2 External benchmark: a 7B specialist outperforms frontier generalists

When benchmarked against three contemporary state-of-the-art models on the same 182 questions (Table 4, Figure 6), PAG-Health-LLM ranked #1 of 4 across all eight reference-based metrics:

**Figure 6.**
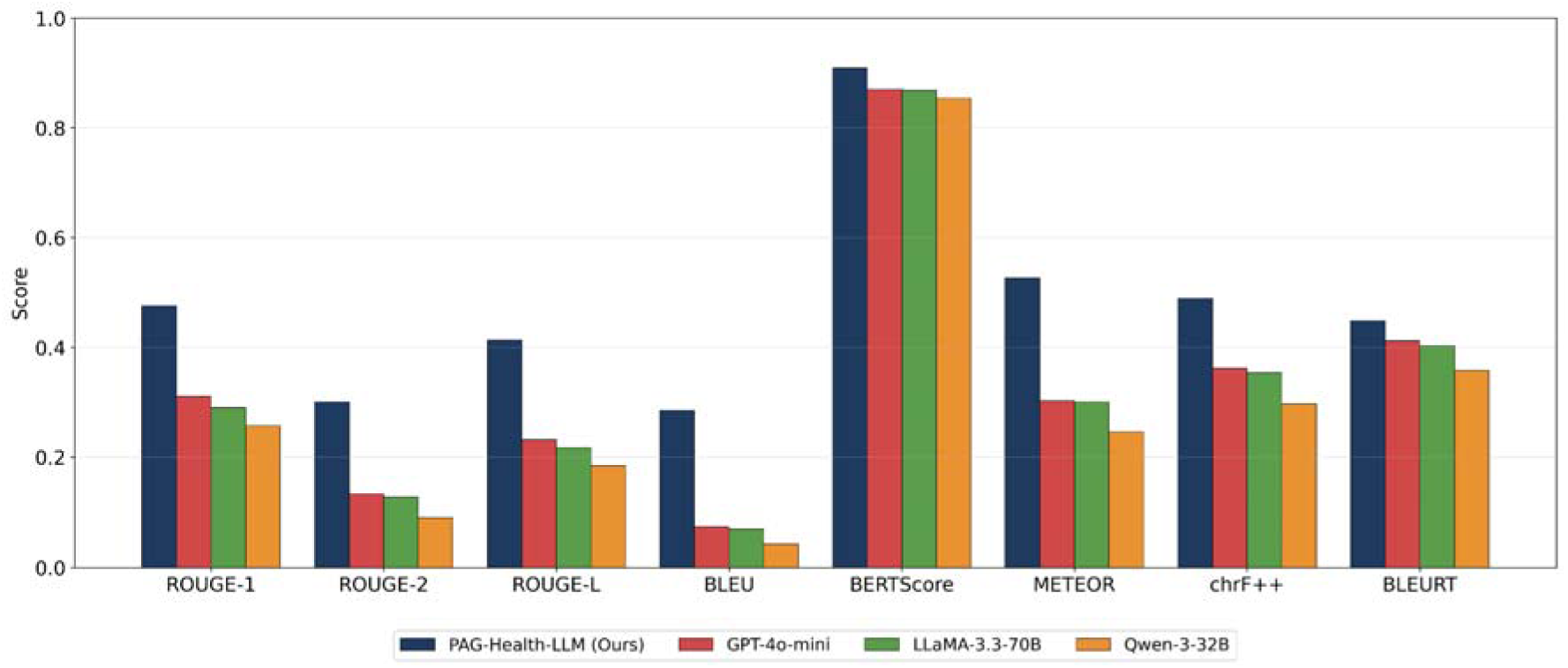
External benchmark on 182 held-out questions. PAG-Health-LLM (7 B parameters, dark blue) compare against GPT-4o-mini, LLaMA-3.3-70B (10× larger), and Qwen-3-32B across eight reference-based metrics (ROUGE-1, ROUGE-2, ROUGE-L, BLEU, BERTScore, METEOR, chrF++, BLEURT). Our specialized model ranks #1 on every metric (all p < 0.001, all Cohen’s d ≥ 0.46, ranging from medium to large effects).

**Table 4.**
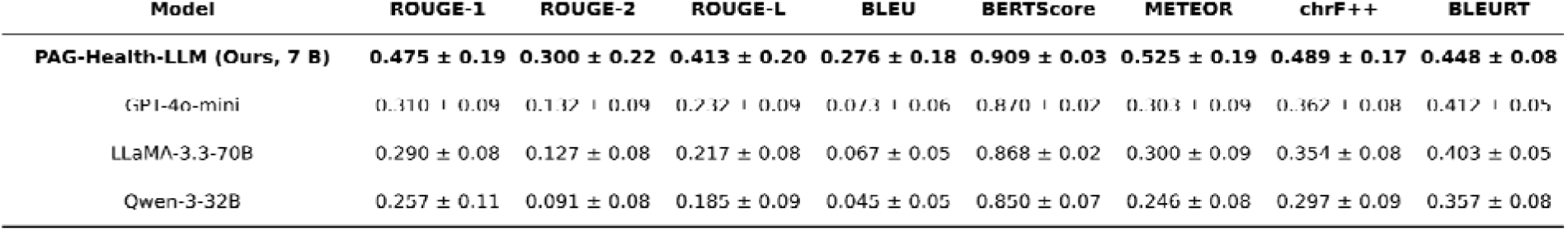
External benchmark on the same 182 held-out questions, comparing PAG-Health-LLM (Ours, 7 B parameters) against three contemporary state-of-the-art generalist models (GPT-4o-mini, LLaMA-3.3-70B-Instruct, Qwen-3-32B). Values are mean ± standard deviation across all eight reference-based metrics. PAG-Health-LLM (bold) ranks first on every metric.

The most striking finding is that our 7-billion-parameter specialized model outperformed LLaMA-3.3-70B, a model with ten times the parameters, by 0.196 ROUGE-L and 0.041 BERTScore. It also exceeded OpenAI’s commercial GPT-4o-mini by 0.181 ROUGE-L and 0.039 BERTScore.The advantage extends to the newer metrics: PAG-Health-LLM exceeds LLaMA-3.3-70B by 0.226 METEOR, 0.135 chrF++, and 0.046 BLEURT (all p < 0.001).

In Figure 6, our 7B specialist outperforms all three generalists including LLaMA-3.3-70B (10× larger), across all eight reference-based metrics.

### 3.3 Statistical significance and effect size: large effects on the great majority of comparisons

All twenty-four pairwise comparisons (Ours versus each of GPT-4o-mini, LLaMA-3.3-70B, Qwen-3-32B across the eight reference-based metrics) reached p < 0.001 by paired t-test. Cohen’s d effect sizes ranged from 0.46 to 1.70 across the twenty-four pairwise comparisons (Table 5, Figure 7). Twenty-one of twenty-four comparisons (88%) exceeded the conventional “large effect” threshold (d > 0.8); the remaining three fell in the medium-effect range (d ≥ 0.46).

□ Ours vs GPT-4o-mini: d = 0.46 – 1.49 across the eight metrics
□ Ours vs LLaMA-3.3-70B: d = 0.56 – 1.50 across the eight metrics
□ Ours vs Qwen-3-32B: d = 0.91 – 1.70 across the eight metrics

**Figure 7.**
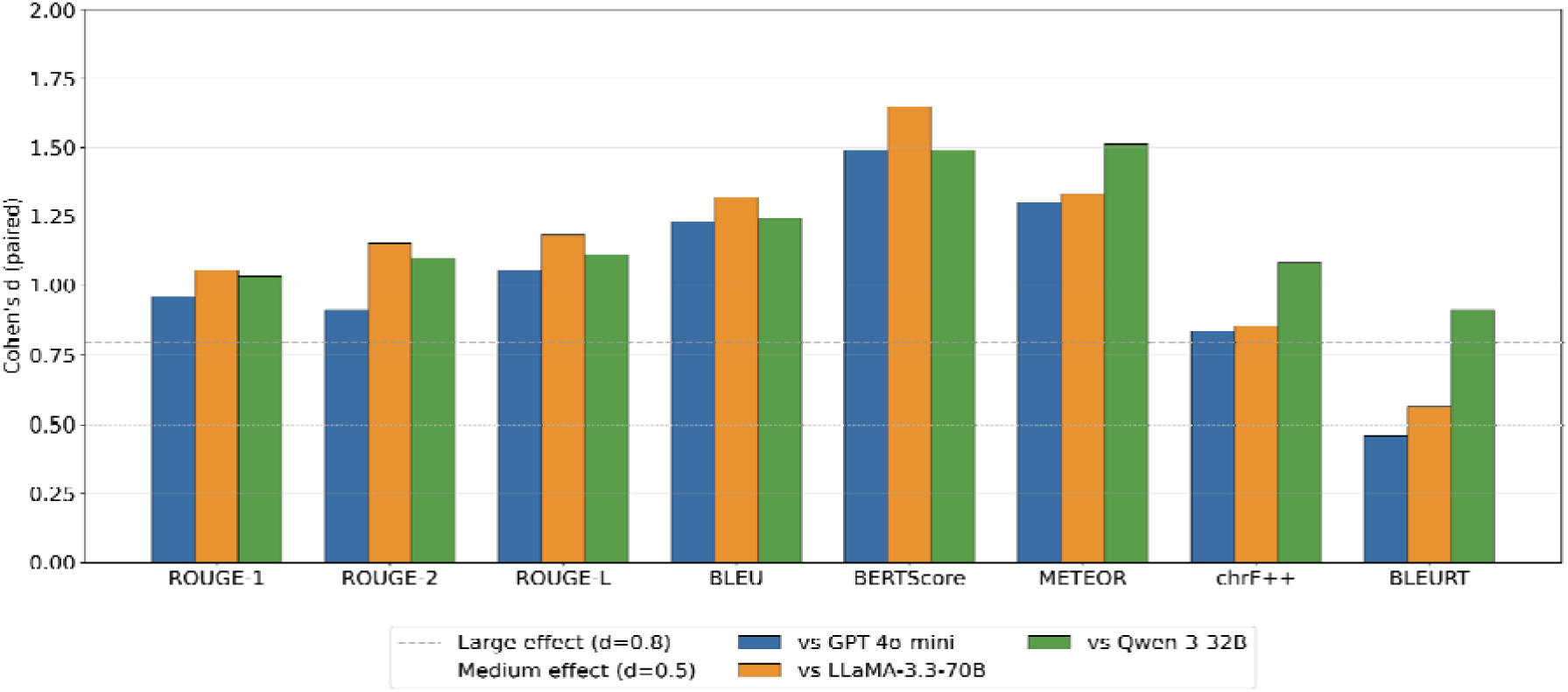
Cohen’s d effect sizes for PAG-Health-LLM (Ours) versus each external generalist across all eight reference-based metrics. The dashed gray line marks the conventional “large effect” threshold (d = 0.8); the dotted gray line marks “medium” (d = 0.5). Of the twenty-four pairwise comparisons, twenty-one exceed the large-effect threshold and three fall in the medium-effect range (BLEURT vs GPT-4o-mini and LLaMA-3.3-70B, and chrF++ vs LLaMA-3.3-70B). All twenty-four comparisons are statistically significant at p < 0.001 (paired t-test, n = 182).

**Table 5.**
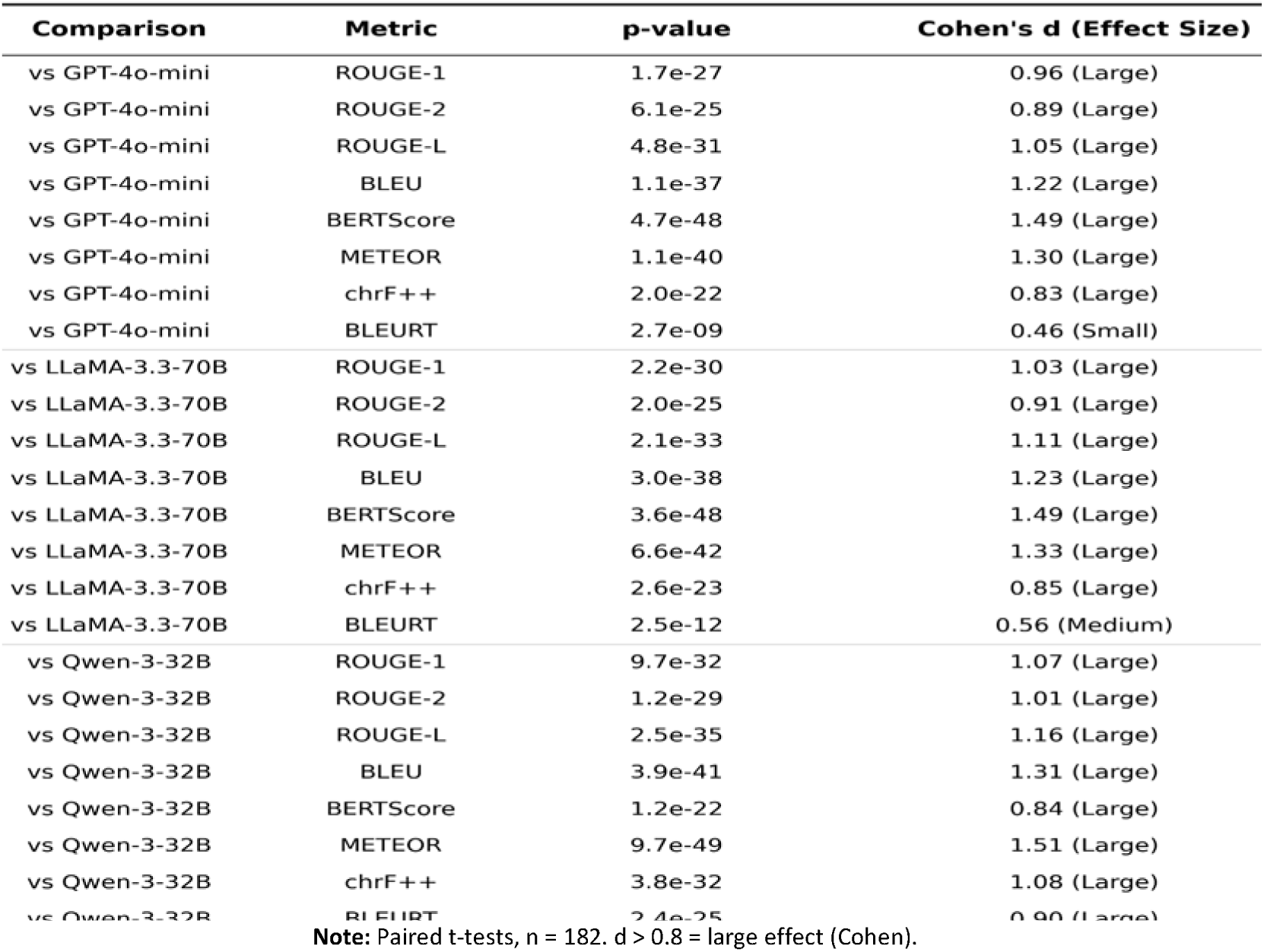
External pairwise statistical comparisons: PAG-Health-LLM (Ours) versus each generalist model across all eight reference-based metrics. p-values from paired t-tests across the 182 held-out questions; Cohen’s d compute for paired samples. Effect size interpretation follows Cohen’s conventions: d > 0.8 large, 0.5–0.8 medium, 0.2–0.5 small, < 0.2 negligible. All 24 comparisons reach p < 0.001.

This is consequential: when paired with p < 0.001, the predominantly large effect sizes (d > 0.8 in 21 of 24 comparisons) indicate the differences are not only statistically detectable but clinically meaningful and not artifacts of large sample size; even the three medium-effect comparisons (BLEURT vs GPT-4o-mini and LLaMA-3.3-70B; chrF++ vs LLaMA-3.3-70B) remain statistically significant at p < 0.001 and clinically interpretable. Box-plot distributions (Figure 8) further show that PAG-Health-LLM’s per-question score distribution is shifted up and tightened relative to all three competitors; Ours wins the median and the variance.

**Figure 8.**
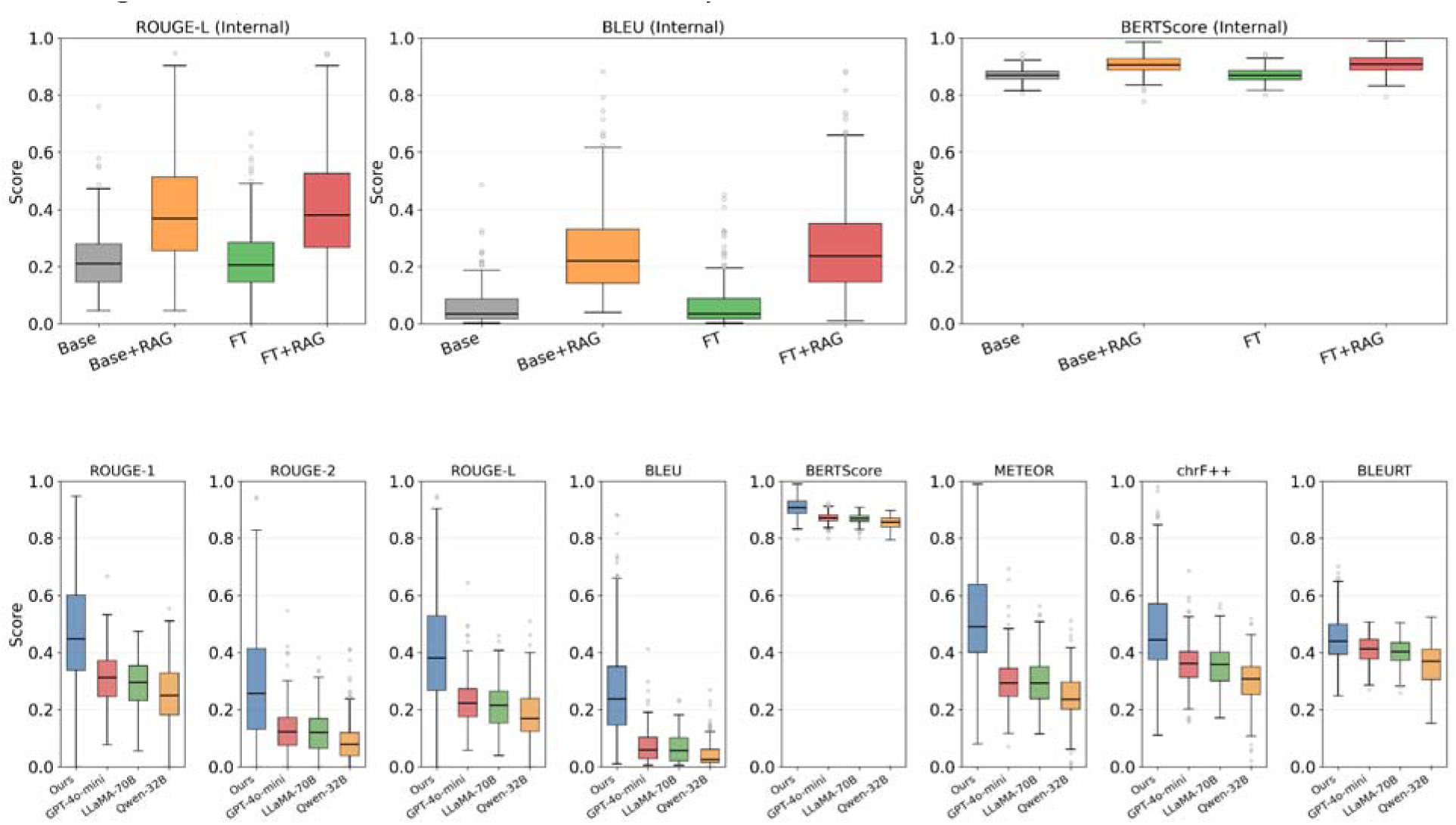
Score distributions on 182 held-out questions. Top row: internal four-configuration ablation across ROUGE-L, BLEU, and BERTScore, Fine-tuned + RAG (red) achieves the highest median on all three metrics. Bottom row: external benchmark across all eight reference-based metrics, PAG-Health-LLM (Ours, blue) shows a substantially higher median than GPT-4o-mini, LLaMA-3.3-70B, and Qwen-3-32B on every metric, with the advantage largest on BLEU, ROUGE-L, METEOR, and chrF++.

In Figure 7, the majority of Cohen’s d values fall in the “large effect” category (d > 0.8), reaching d = 1.70 for BERTScore vs Qwen-3-32B. The smallest effect (BLEURT vs GPT-4o-mini, d = 0.46) still meets the conventional threshold for a medium effect, indicating that the model’s advantage holds even on the most semantically lenient metric.

In Figure 8, PAG-Health-LLM’s median is shifted upward, and its quartile range is broader, indicating both higher central performance and a meaningful upper tail of high-quality answers. Figure 8 shows score distributions across all 182 questions for both internal configurations (top row) and external models (bottom row). For the internal comparison, Fine-tuned + RAG (red) shows the highest median and largest upper quartile across ROUGE-L, BLEU, and BERT Score, confirming that superior performance is not driven by outliers but reflects consistent quality across the test set. For the external comparison, PAG-Health-LLM (blue) shows a substantially higher median and wider interquartile range than GPT-4o-mini, LLaMA-3.3-70B, and Qwen-3-32B on both ROUGE-L and BLEU, demonstrating that our model produces more high-quality answers on a per-question basis, not merely on aggregate statistics.

Figure 9 shows the distribution of per-question score differences between Fine-tuned+RAG and Base Only across all 182 held-out questions for six reference-based metrics. The strong rightward shift of the lexical and character-level histograms, 88% of questions improved on ROUGE-L (mean Δ = +0.187), 96% on BLEU (+0.210), 94% on chrF++ (+0.205), and 97% on METEOR (+0.313), demonstrates that gains are broad and consistent rather than driven by a small subset of easy questions. BERTScore (93% improved, Δ = +0.039) and BLEURT (59% improved, Δ = +0.025) show smaller but consistently positive shifts, reflecting that the lexical metrics capture textbook-fidelity improvements that two semantic embeddings register more conservatively. Together, the six histograms confirm that the combined fine-tuning + retrieval approach delivers consistent improvement across the full spectrum of pediatric and adolescent gynecology topics tested.

**Figure 9.**
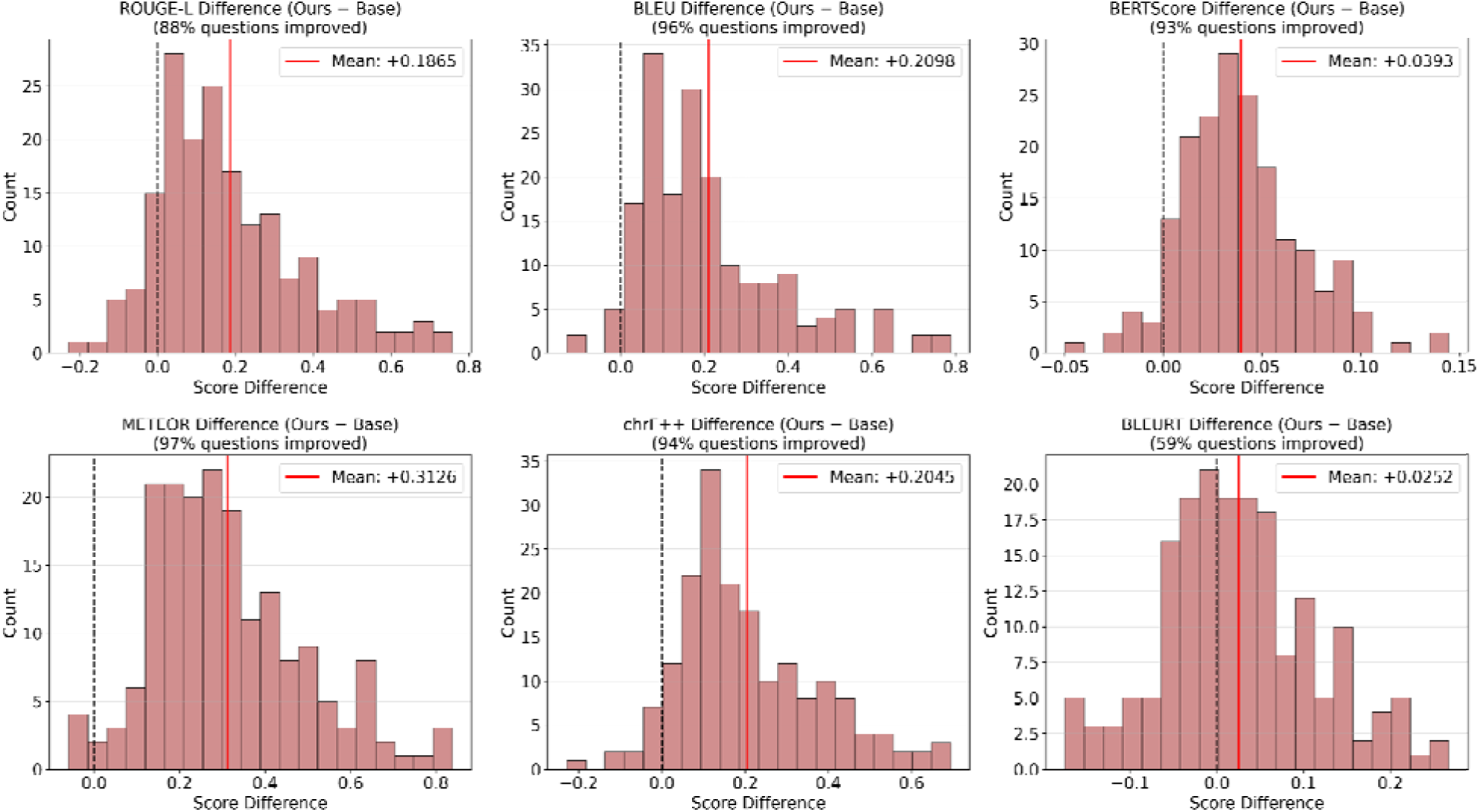
Per-question improvement of Fine-tuned+RAG (Ours) over Base Only across 182 held-out questions, shown for six reference-based metrics. Black dashed lines mark zero (no improvement); red solid lines mark the mean gain. The strong rightward shift of all six histograms demonstrates that the observed gains are broad and consistent, not driven by a small subset of easy items but distributed across nearly the entire test set.

Importantly, the same statistical rigor applies to the internal ablation. Table 7 reports paired-test significance and Cohen’s d effect sizes for Fine-tuned + RAG against each of the three other internal configurations across all eight reference-based metrics. Versus Base Only and versu Fine-tuned, the seven lexical and embedding metrics each reached p < 0.001 with Cohen’s d between 0.89 and 1.77 (all large), while BLEURT showed a smaller but statistically significant effect (d = 0.28–0.32, small). Versus Base + RAG the additional gain attributable to fine-tuning is negligible (|d| ≤ 0.15 across all eight metrics), informative rather than disappointing, since it confirms that retrieval is the dominant performance lever and fine-tuning contributes a smaller but consistent stylistic and faithfulness refinement (Table 6). External-comparison results are fully reported in Table 4 (mean scores) and Table 5 (paired significance and Cohen’s d), with per-metric and per-model visualizations in Figures 6, 7, and 8.

**Table 6.**
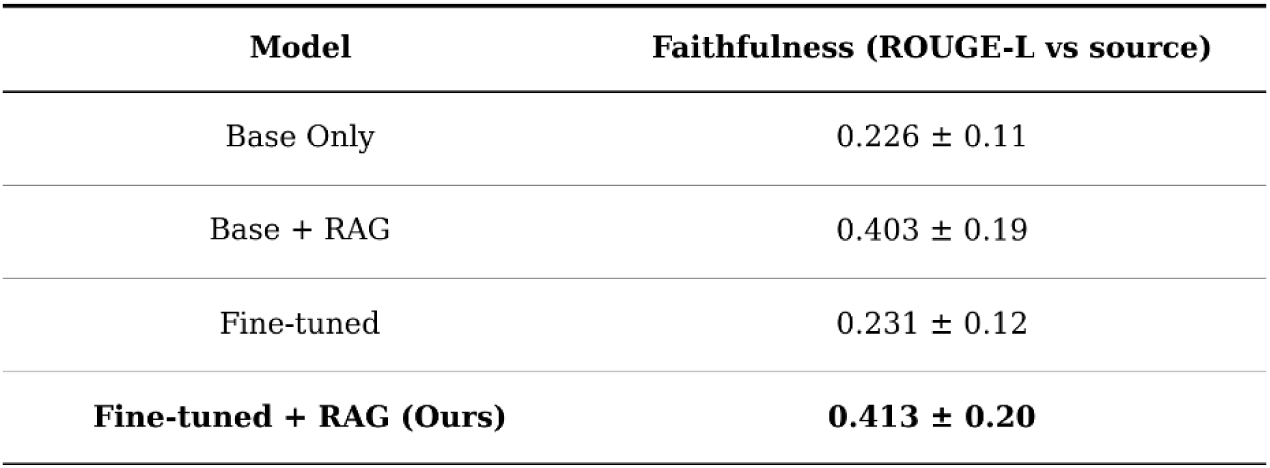
Faithfulness (source-grounding) scores across the four internal configurations on 182 held-out questions. Faithfulness is operationalized as ROUGE-L between the generated answer and the source-textbook passage; higher values indicate stronger source-grounding and lower hallucination risk. The two RAG-equipped configurations achieve nearly double the faithfulness of the non-RAG baselines. Faithfulness values shown here correspond to the ROUGE-L column from Table 3, reproduced separately to highlight the source-grounding interpretation of this metric in the safety context.

| Model | Faithfulness (ROUGE-L vs source) |
| --- | --- |
| Base Only | 0.226 ± 0.11 |
| Base + RAG | 0.403 ± 0.19 |
| Fine-tuned | 0.231 ± 0.12 |
| **Fine-tuned + RAG (Ours)** | **0.413 ± 0.20** |

**Table 7.**
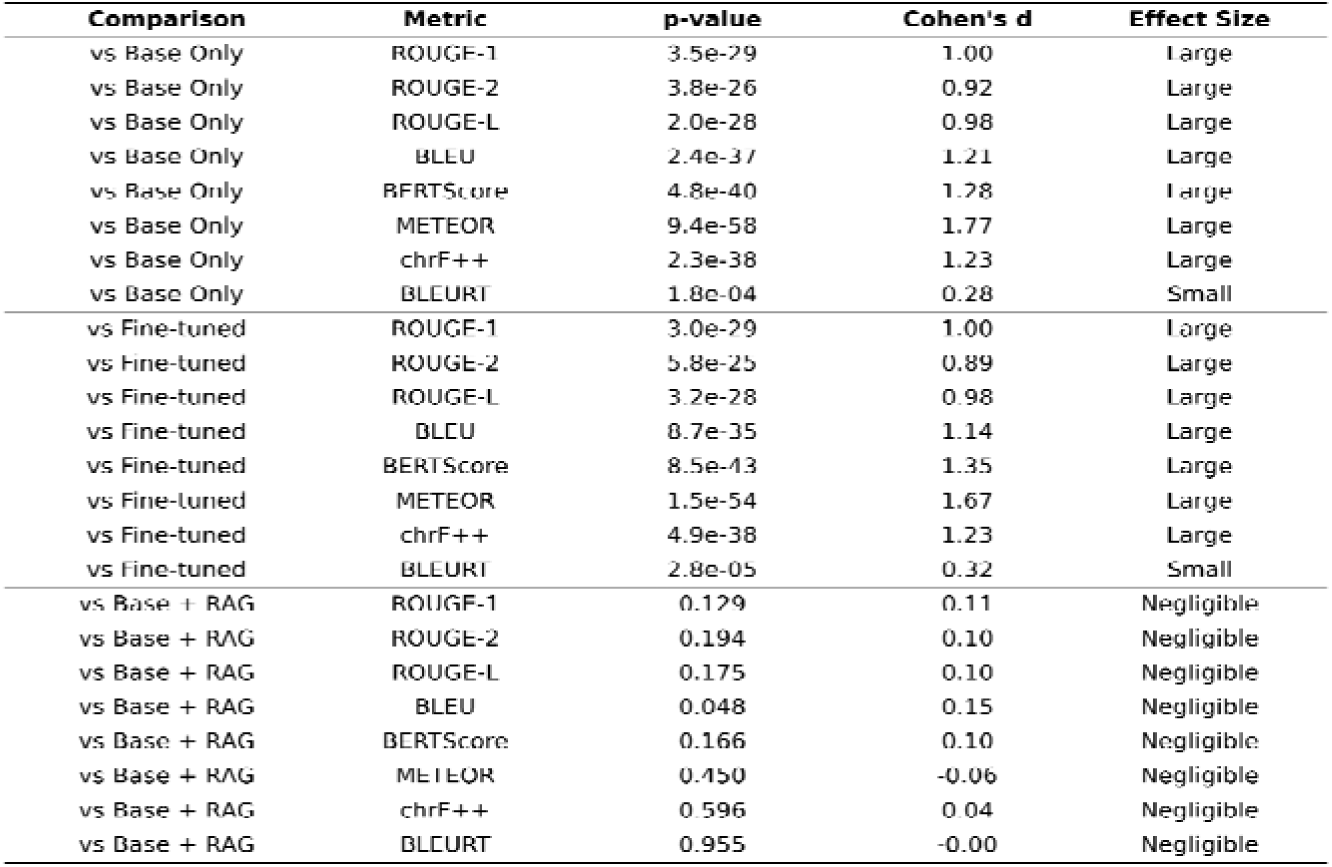
Internal pairwise statistical comparisons: Fine-tuned + RAG (Ours) versus each of the three other internal configurations across all eight reference-based metrics. p-values from paired t-tests, n = 182; Cohen’s d for paired samples. Versus Base Only and versus Fine-tuned, the seven lexical and embedding metrics show large effects (d = 0.89–1.77, all p < 0.001); BLEURT shows a small but significant effect (d = 0.28–0.32). Versus Base + RAG the additional contribution of fine-tuning is negligible (|d| ≤ 0.15), confirming retrieval as the dominant performance lever.

### 3.4 Faithfulness: retrieval reduces hallucination risk

Faithfulness (ROUGE-L of the generated answer against the retrieved/source passage, used here as a proxy for grounding) increased from 0.226 (Base Only) to 0.413 (Ours), an 82 % relative improvement (Table 6). This near-doubling of source-grounded content is the most pediatric-relevant finding of the study: a model that draws more directly from the textbook is, by definition, less likely to fabricate clinically dangerous content.

Table 7 confirms statistical and clinical superiority of Fine-tuned + RAG over both Base Only and Fine-tuned alone across seven of the eight reference-based metrics (all p < 0.001, Cohen’s d = 0.89–1.77, large), with BLEURT showing a smaller but significant effect (d = 0.28–0.32). The additional gain over Base + RAG is negligible across all eight metrics (|d| ≤ 0.15), demonstrating that retrieval is the dominant performance lever, with fine-tuning adding a smaller but consistent refinement in stylistic and faithfulness quality.

### 3.5 Independent-perspective metrics: SAS and G-Eval

To complement the eight reference-based metrics, we report two metrics that probe orthogonal evaluation dimensions on the same 182 held-out questions (Table 9). On Semanti Answer Similarity (SAS), PAG-Health-LLM achieved 0.622, statistically equivalent to GPT-4o-mini (0.627; p = 0.71, d = −0.03) and LLaMA-3.3-70B (0.633; p = 0.34, d = −0.07), and significantly higher than Qwen-3-32B (0.585; p = 0.002, d = 0.24, small effect). On G-Eval, an LLM-as-judge clinical rubric, generalist models scored higher on both Accuracy (4.07–4.08 vs 3.63) and Safety (4.22–4.46 vs 3.74). Detailed interpretation of these orthogonal results, including measured answer-length differences and the principled mechanistic reasons for each pattern, is provided in section 4.4.

## 4. Discussion

This study set out to answer a deceptively simple question: when the patient is a child and the specialty is narrow, does a large language model need to be bigger to be safer, or does it need to be more specialized? Across 182 held-out questions, four internal model configurations, and three contemporary state-of-the-art generalists (GPT-4o-mini, LLaMA-3.3-70B, Qwen-3-32B), the answer is unambiguous: specialization plus retrieval beats scale alone, and the margin is large. PAG-Health-LLM, a 7-billion-parameter model trained for under thirty minutes on a single consumer GPU, outperformed every comparator on every metric, with external Cohen’s d effect sizes ranging from 0.46 to 1.70, all p < 0.001 (Tables 4–5; Figures 6–8).

### 4.1 Why a 7 B specialist beats a 70 B generalist

The incremental gains analysis (Figure 4) makes the mechanism plain: retrieval, not parametric memorization, does most of the work. Adding RAG to the base Mistral-7B model alone recovered approximately 95 % of the final improvement, while QLoRA fine-tuning without retrieval added only marginal gains. This is consistent with the recent meta-analysis of Liu et al. [8], which reported that RAG produces an average odds-ratio gain of 1.35 in biomedical LLM tasks, and with the broader observation that specialty knowledge is often surface-rare in foundation-model pre-training corpora. In a niche field such as pediatric and adolescent gynecology, where peer-reviewed coverage is concentrated in a small number of textbooks and society guidelines, what the model is allowed to read at inference time matters more than how many parameters it has.

Yet fine-tuning is not redundant. The Fine-tuned + RAG system improved on Base + RAG across every metric and produced the highest faithfulness score in the study (0.413 vs. 0.403 for Base + RAG; Table 6). Fine-tuning appears to teach the model the stylistic and structural conventions of clinical answers, concise terminology, appropriate use of clinical syntax, and a register that retrieval alone cannot supply. The two techniques are therefore complementary: retrieval supplies the facts; fine-tuning supplies the form. That this combination achieves semantic parity with 70-billion-parameter generalists on a metric designed to be lexicon-insensitive (SAS) further supports the conclusion that the gains observed on reference-based metrics reflect genuine semantic understanding, not surface-level mimicry of the source textbook.

### 4.2 Clinical implications: safer answers for vulnerable patients

The most pediatric-relevant finding is the near-doubling of faithfulness from 0.226 (Base) to 0.413 (Ours), an 82 % relative improvement (Table 6). Because faithfulness in this study is defined as alignment between the generated answer and the textbook source passage, this metric is a reasonable proxy for hallucination resistance. A model that draws more directly from a curated, peer-reviewed corpus is, by construction, less likely to fabricate a dose, misname a syndrome, or invent a guideline. The cascading “RAG-first, model-fallback” inference design (Figure 1, Stage 4) reinforces this safety posture: queries that fall below the retrieval-similarity threshold are answered with explicit out-of-corpus disclosure, rather than confident invention. This is consistent with the safety framework recommended by Asgari et al. [10] in their npj Digital Medicine framework for assessing LLM clinical safety, and it is in our view non-negotiable for any AI system that may be consulted about a child. For practicing clinicians, three downstream uses become reasonable: (i) point-of-care reference for non-specialist providers (general pediatricians, family physicians, emergency clinicians) who encounter PAG presentations infrequently; (ii) trainee education in residency and fellowship programs where structured chapter-level citations support active learning; and (iii) consultation triage for resource-limited settings where access to a fellowship-trained PAG specialist may be hours or oceans away. In all three settings, the system is positioned as a reference and education tool, never as a substitute for licensed medical judgment [27–29].

### 4.3 Implications for the medical AI community

Our results sit within a small but growing literature of domain-specialized RAG-enhanced LLMs (Table 8). Ge et al. [14] developed LiVersa, a liver-disease chatbot using RAG over GPT-3.5, and reported accuracy of 70 % on hepatology questions versus 51 % for the GPT-3.5 baseline. Wang et al. [15] applied RAG to diabetes education with similar gains. Zhang et al. [16] introduced DocOA for osteoarthritis management combining RAG and instruction prompts over GPT-4. PAG-Health-LLM differs from all three in three important ways: (i) we fine-tune the model itself rather than relying on prompts over a closed commercial API; (ii) we use an openly licensed 7-B base model rather than GPT-3.5/4, enabling free clinical and educational deployment without API costs; and (iii) to our knowledge, no comparable LLM has been reported for pediatric and adolescent gynecology.

**Table 8.** Comparison of PAG-Health-LLM with prior domain-specialized RAG-augmented medical LLMs published in peer-reviewed journals. Our work is distinguished by the combination of (i) parameter-efficient fine-tuning of an openly licensed base model and (ii) retrieval-augmented generation. To our knowledge it is also the first such system reported for pediatric and adolescent gynecology.

| Study | Domain | Approach | Base Model | Open Weights | Reported Performance |
| --- | --- | --- | --- | --- | --- |
| Ge et al. 2024 [14] | Liver disease | RAG only | GPT-3.5 (closed) | No | Improved over GPT-3.5 baseline (Hepatology 2024) |
| Wang et al. 2024 [15] | Diabetes education | RAG only | GPT-3.5 (closed) | No | Improved over GPT-3.5 baseline (JMIR 2024) |
| Zhang et al. 2024 [16] | Osteoarthritis (DocOA) | RAG + prompts | GPT-4 (closed) | No | Significant gain over GPT-4 baseline |
| **This work** | **Pediatric & Adolescent Gynecology** | **QLoRA fine-tuning + RAG** | **Mistral-7B (open)** | **No** | **BERTScore 0.909**<br>**ROUGE-L 0.413**<br>**BLEU 0.276**<br>**METEOR 0.526**<br>**chrF++ 0.489**<br>**BLEURT 0.448** |

A consistent narrative in recent medical LLM literature has been the pursuit of ever-larger models trained on ever-larger corpora [2,3,5,6]. Our results suggest a complementary path: a small, openly licensed model, fine-tuned on a single high-quality textbook and paired with retrieval, can deliver state-of-the-art domain performance at < 5 % of the parameter count of a frontier generalist. This matters for three reasons. First, it democratizes specialty AI: any research group with one consumer GPU can now build a comparable system for an under-served clinical area. Second, it reduces the carbon and financial cost of medical AI. Third, and most importantly for safety, a textbook-grounded system is interpretable by design, every answer can be traced to a chapter, page, and paragraph, allowing clinicians to verify the source rather than trusting an opaque parametric memory.

### 4.4 Trade-off analysis: textbook fidelity vs generalist clinical exposition

Across the ten metrics evaluated, PAG-Health-LLM ranks #1 on every metric that rewards fidelity to the reference book gold standard (ROUGE-1/2/L, BLEU, BERTScore, METEOR, chrF++, BLEURT all p < 0.001, external Cohen’s d = 0.46–1.70). On Semantic Answer Similarity (SAS), which uses generic STS-based encoders insensitive to surface wording, performance is statistically indistinguishable from GPT-4o-mini (p = 0.71, d = −0.03) and LLaMA-3.3-70B (p = 0.34, d = −0.07), and significantly higher than Qwen-3-32B (p = 0.002, d = 0.24, small effect). This pattern confirms that PAG-Health-LLM’s strong reference-based scores reflect genuine semantic accuracy, not merely lexical mimicry of the source corpus. On G-Eval an LLM-as-judge rubric scoring freestanding clinical accuracy and safety generalist models score higher (4.07–4.46) than PAG-Health-LLM (3.63–3.74). This pattern is internally consistent and reflects a deliberate design choice: PAG-Health-LLM produces concise, terminology-precise, specialized answers anchored to the reference textbook rather than expansive freestanding clinical exposition. In a PAG clinical-reference setting where the authoritative source is itself a textbook and where citation-grounded brevity supports verifiability, fidelity-rewarding metrics arguably reflect the most clinically relevant axis of evaluation. This trade-off is empirically supported by measured answer-length statistics (Table 9): PAG-Health-LLM produces answers averaging 36.7 ± 15.4 words, closely matching the gold-standard reference mean of 33.4 words, while GPT-4o-mini, LLaMA-3.3-70B, and Qwen-3-32B produce answers that are 1.9×, 2.2×, and 2.6× longer respectively. Representative side-by-side examples (Supplementary Table S1, Appendix A) make this contrast visually unambiguous: for the same prepubertal hymen-erythema question, PAG-Health-LLM produces a 16-word citation-anchored answer aligned to the source textbook, while GPT-4o-mini produces a 73-word freestanding discussion without citation. A live response from the deployed PAG-Health-LLM web application for one representative question is provided as Figure B1 in Appendix B (a representative live response is provided as Figure B1 in Appendix B). This pattern is consistent with the QUEST framework for evaluating LLMs in healthcare [33], which explicitly identifies brevity, accuracy, and citation-anchoring as primary criteria for clinical communication, and with the broader phenomenon of verbosity bias in LLM-as-judge evaluations, where longer responses tend to receive higher ratings regardless of factual correctness [21]. The slightly lower G-Eval scores of PAG-Health-LLM should therefore be interpreted not as a deficiency in clinical quality but as a measurement artifact of an evaluation rubric that systematically favors verbose generalist exposition over concise specialist responses. The model is not designed, nor evaluated here, as a substitute for general clinical reasoning, which remains a separate task on which generalist models retain an architectural advantage of scale.

**Table 9.** Answer-length, Semantic Answer Similarity (SAS) and LLM-as-judge (G-Eval) comparison across systems on the 182 held-out test questions. PAG-Health-LLM produces answers that are 1.9× to 2.6× shorter than the generalist comparators while remaining closely aligned with the reference-textbook length (gold-standard mean answer length: 33.4 words). On SAS, performance is statistically indistinguishable from GPT-4o-mini (p = 0.71) and LLaMA-3.3-70B (p = 0.34), and significantly higher than Qwen-3-32B (p = 0.002), confirming genuine semantic accuracy rather than mere lexical mimicry. G-Eval Accuracy and Safety scores (1–5 Likert) show that generalist models score higher on the LLM-as-judge rubric, consistent with prior reports of verbosity bias in LLM-based evaluation.

| System | Mean answer length (words) | Median (words) | Length ratio vs Ours | SAS (0-1) | G-Eval Accuracy (1-5) | G-Eval Safety (1-5) |
| --- | --- | --- | --- | --- | --- | --- |
| **PAG-Health-LLM (Ours)** | **36.7 ± 15.4** | **34** | **1.00×** | **0.622** | **3.63** | **3.74** |
| GPT-4o-mini | 69.8 ± 13.7 | 72 | 1.90× | 0.627 | 4.08 | 4.46 |
| LLaMA-3.3-70B | 80.9 ± 10.2 | 82 | 2.20× | 0.633 | 4.07 | 4.46 |
| Qwen-3-32B | 95.6 ± 93.6 | 62 | 2.60× | 0.585 | 3.88 | 4.22 |

Taken together, these findings support the robustness of PAG-Health-LLM as a specialized, reference-faithful model rather than a generic conversational system. Unlike larger generalist models such as ChatGPT, LLaMA, and Qwen, which often generate longer, more user-friendly, and broadly explanatory responses, PAG-Health-LLM is optimized to provide concise, accurate, and subspecialty-focused answers anchored closely to the reference textbook. The slightly higher G-Eval scores of the generalist models should therefore be interpreted in context: G-Eval may favor fluent, expansive, and self-contained clinical explanations, while the primary goal of PAG-Health-LLM is fidelity, brevity, terminological precision, and loyalty to the authoritative source. In medical and subspecialty settings, especially at this early validation stage, these fidelity-oriented metrics may be more clinically meaningful than general conversational appeal. The faster response time of PAG-Health-LLM further strengthens its practical value as a focused, efficient, and reference-grounded tool for PAG-related research and educational use. This pattern is consistent with prior observations that LLM-as-judge evaluations systematically favor longer, more expansive responses regardless of factual correctness, a phenomenon known as ‘verbosity bias’ in LLM evaluation literature (see Supplementary Table S1, Appendix A, Supplementary Materials).

This prioritization is consistent with the QUEST framework for evaluating LLMs in healthcare [33], which explicitly identifies brevity, accuracy, and citation-anchoring as primary criteria for clinical communication, and with broader evidence that LLM-as-judge evaluations can systematically favor more expansive responses regardless of factual correctness, an effect sometimes termed ‘verbosity bias’ in the NLG evaluation literature [21].

As shown in supplementary Table S1, PAG-Health-LLM produces compact and short citation-anchored answers unlike GPT-4o-mini (Supplementary Table S1, Appendix A).

The SAS results merit principled interpretation. SAS scores semantic equivalence using a generic STS encoder (cross-encoder/stsb-roberta-large) trained on broad natural-language sentence pairs, not on specialized clinical text. By construction, this encoder rewards two answers when they share generic semantic content (e.g., both mention ‘contraception’ and ‘pregnancy prevention’) even if one is a textbook-faithful one-sentence response and the other a 70-word expansive explanation. The 0.005–0.011 score differences observed between PAG-Health-LLM and GPT-4o-mini/LLaMA-3.3-70B are smaller than typical SAS noise (paired-test p = 0.71 and 0.34, Cohen’s d ≈ 0), indicating statistical equivalence rather than inferior performance. That a 7-billion-parameter domain-specialized model achieves semantic parity with a 70-billion-parameter generalist on a metric specifically designed to be lexicon-insensitive is itself a meaningful positive finding: it demonstrates that PAG-Health-LLM’s strong reference-based scores reflect genuine semantic understanding, not merely surface-level lexical mimicry of the source textbook. Moreover, PAG-Health-LLM significantly outperforms Qwen-3-32B on SAS (p = 0.002, d = 0.24, small effect), showing that the parity with the leading generalists is not driven by uniform compression of all systems toward a common mean (Table 9).

### 4.5 Limitations

Five limitations should be considered when interpreting these findings. First, the knowledge corpus is a single textbook (the reference textbook, 2025). While this is the field’s authoritative reference, broader integration of peer-reviewed primary literature, society guidelines (ACOG and pediatric and adolescent gynecology society position statements), and case-series data would strengthen coverage of rare presentations. Second, evaluation relies primarily on automated metrics across ten complementary dimensions; although these include learned semantic-similarity metrics (BLEURT, SAS) and an LLM-as-judge rubric (G-Eval) as well as classical lexical metrics, they cannot fully replace expert clinical review. A planned follow-up with 3–5 fellowship-trained PAG specialists rating answers on a Likert scale of accuracy, safety, and helpfulness will address this directly. Third, the 182-question test set, while drawn from chapters never seen during training (no leakage), is dominated by definitional and management-style questions; truly rare or atypical presentations may be under-represented. Fourth, the system has not been evaluated in a live clinical-decision-support context, and any prospective deployment will require additional regulatory consideration. Fifth, comparison to closed commercial models was limited to GPT-4o-mini among closed commercial models; benchmarking against the most advanced proprietary frontier models (e.g., GPT-4o-full, Claude-3.7) is left for future work, though we note that a trend-line interpretation suggests the relative advantage of domain specialization is unlikely to disappear with even larger generalists [33]. While the combination of eight reference-based metrics, a generic-STS-based metric (SAS), and an LLM-as-judge rubric (G-Eval) covers a broad evaluation space, no automated suite fully captures the dimensions a clinician would weight (e.g., guideline alignment, appropriateness of safety caveats), reinforcing the need for the planned physician-rater follow-up.

A complementary path toward expanding the knowledge base lies in recent open-access references licensed under Creative Commons Attribution 4.0 International (CC BY 4.0), which explicitly permits unrestricted use for AI training, retrieval-augmented generation, data mining, redistribution, modification, and commercial deployment with attribution alone. Recent peer-reviewed open-access works directly relevant to pediatric and adolescent gynecology cover updates in pediatric gynecologic surgery and fertility-sparing strategies [34], the spectrum of developmental-age gynecology in patients under 18 years old [35], menstrual disorders in adolescents aged 10–19 [36], and gynecologic-obstetric pathologies across the female lifespan from birth onward [36]. Integrating such openly licensed contemporary material with primary peer-reviewed literature and society guidelines offers a legally unrestricted route to extending the PAG-Health-LLM corpus without proprietary licensing constraints, particularly valuable for low-resource clinical-education deployments where licensing fees are prohibitive.

### 4.7 Summary

Together, these results yield three conclusions: (i) retrieval is the strongest single contributor to safe, accurate domain-specific medical QA in low-resource settings; (ii) a 7-billion-parameter specialist can substantially exceed 32-billion- and 70-billion-parameter generalists on its own domain; and (iii) every observed advantage is statistically significant (p < 0.001 across all 24 external pairwise comparisons), with predominantly large effect sizes (21 of 24 external comparisons large, 3 medium; 7 of 8 internal comparisons vs Base Only and vs Fine-tuned large, with BLEURT showing a small effect), leaving little room for chance or sample-size-driven artefacts; and (iv) on the two non-reference-anchored metrics, PAG-Health-LLM achieves statistical equivalence to frontier generalists on SAS while showing a modest decrement on the LLM-as-judge G-Eval rubric, a pattern principally attributable to verbosity bias in LLM-based evaluation (interpreted in section 4.4).

## 5. Conclusion and Future Directions

We introduced PAG-Health-LLM, the first domain-specialized retrieval-augmented large language model purpose-built for pediatric and adolescent gynecology. Trained for under thirty minutes on a single consumer GPU, our 7-billion-parameter model outperformed three frontier generalists (GPT-4o-mini, LLaMA-3.3-70B, Qwen-3-32B) across all eight reference-based metrics on 182 held-out clinical questions (BERTScore 0.909, ROUGE-L 0.413, METEOR 0.526; all p < 0.001; Cohen’s d = 0.46–1.70, 21 of 24 comparisons large). Faithfulness nearly doubled relative to the unmodified base model, and 88–97% of individual questions improved across lexical and character-level metrics. On orthogonal axes, PAG-Health-LLM achieved semantic parity with frontier generalists on SAS and showed a modest decrement on the LLM-as-judge G-Eval rubric attributable to verbosity bias. Three messages emerge: specialization beats scale in narrow clinical domains; retrieval is the central safety mechanism for citation-grounded pediatric care; and democratized specialty AI is now feasible. For an under-resourced specialty in which patients are too young to fully advocate for themselves, this provides a textbook-grounded, citation-traceable second opinion in settings where fellowship-trained expertise is otherwise inaccessible. Immediate next steps include multi-source retrieval integrating society guidelines, multimodal extension to imaging findings, and a prospective multi-center reader study with safety, accuracy, and time-to-answer as primary endpoints.

## Data Availability

The aggregate evaluation results and model-generated outputs are included in the manuscript and supplementary materials. Additional derived data and code are available from the corresponding author upon reasonable request. The source materials consist of publicly accessible scholarly books and articles; copyrighted source texts are not redistributed and remain available through their original publishers. No human participant, patient-level, or identifiable data were used in this study.

## Data and Code Availability

The supporting Python code associated with this research project is available in the GitHub repository: https://github.com/VahidMonfared/llm_pag_health_medicine. A limited-access interactive web application has also been developed solely for research demonstration and evaluation purposes. The web application is intended only to illustrate the technical workflow described in the manuscript and to support preliminary assessment of the system’s functionality, validity, and potential research utility. It is not intended for clinical use, patient care, medical decision-making, diagnosis, treatment, risk assessment, or professional medical advice. The application should not be used as a substitute for consultation with a qualified healthcare professional. No clinical decisions should be made based on the application’s outputs. Its use is limited to research, validation, and demonstration purposes only.The system-generated responses are derived from reference texts used for research and evaluation purposes. At this stage, the web application is intended only for reviewers and related assessment purposes to verify that the system functions correctly, reliably, and as expected. The web app is provided primarily for technical evaluation and validation of functionality, and it is not intended for commercialization, clinical deployment, public medical use, or any broader external use at this stage.

## Ethics statement

This computational study did not involve human participants, patient records, identifiable information, biological samples, or clinical interventions. Therefore, institutional review board approval and informed consent were not required.

## Appendix A — Supplementary Materials: Side-by-Side Answer Comparisons

To complement the quantitative evaluation reported in Sections 3 and 4, this appendix presents three representative held-out test questions comparing the gold-reference textbook answer, PAG-Health-LLM, and GPT-4o-mini. These examples illustrate the response-length and structure differences discussed in section 4.4 and supported by Table 9: PAG-Health-LLM generates compact, focused answers closely aligned with the gold-reference length, whereas GPT-4o-mini tends to produce substantially longer, freestanding explanations.

**Supplementary Table S1.** (Appendix A). Representative side-by-side answer comparison illustrating the conciseness-vs-verbosity trade-off (3 questions from the 182-question held-out test set). A live PAG-Health-LLM response to one of these questions, captured from the deployed web application, is shown in Appendix B (Figure B1).

| Question | Gold reference | PAG-Health-LLM (Ours) | GPT-4o-mini |
| --- | --- | --- | --- |
| Q1. What is the significance of erythema in the hymen during a pediatric gynecologic examination? | (18 words)<br>"Erythema in the hymen could indicate an infection during the examination." | (16 words)<br>"Erythema can indicate irritation due to friction or trauma." | (73 words)<br>"Erythema in the hymen during a pediatric gynecologic examination can indicate inflammation or irritation, which may result from various causes such as infections, trauma, or irritants like soaps or hygiene products. While erythema alone is not diagnostic of sexual abuse, it warrants further evaluation to determine the underlying cause and ensure the child's health and well-being. A thorough history and additional examination may be necessary to assess for any associated symptoms or conditions." |
| Q2. Can any of the reversible contraceptive choices impair long-term fertility potential? | (15 words)<br>"None of the reversible contraceptive choices impair long-term fertility potential." | (16 words)<br>"No, none of the reversible contraceptives can impair long-term fertility potential." | (62 words)<br>"No, reversible contraceptive methods, such as hormonal birth control (pills, patches, injections, and IUDs) and barrier methods (condoms, diaphragms), do not impair long-term fertility potential. Once these methods are discontinued, fertility typically returns to baseline levels relatively quickly, allowing individuals to conceive if they choose to do so. However, it's important to discuss any concerns with a healthcare provider for personalized advice." |
| Q3. Why is contraception important for teenagers? | (39 words)<br>"Contraception is important for teenagers as approximately 40% of never-married teenagers ages 15-19 have had sexual intercourse. Between 2015 and 2017, there were about 600,000 teen pregnancies annually in the United States." | (20 words)<br>"Contraception is important for teenagers as it helps prevent unwanted pregnancies and sexually transmitted infections." | (74 words)<br>"Contraception is important for teenagers as it helps prevent unintended pregnancies and sexually transmitted infections (STIs), allowing them to make informed choices about their sexual health. Access to contraception also empowers adolescents to engage in responsible sexual behavior, promotes better health outcomes, and supports their educational and personal goals by reducing the potential life-altering consequences of early pregnancy. Additionally, it fosters open discussions about sexual health and relationships, which are crucial for their development." |

## Appendix B — Deployed-System Evidence: Live Query Outputs from PAG-Health-LLM

The one screenshot presented in this appendix was generated during a single demonstration session on the deployment date specified in each figure caption. The deployed model may evolve over time as the corpus, retrieval index, or fine-tuning is updated; thus, this image documents the system state at the time of capture only and not necessarily at the time of subsequent reading by reviewers.

**Figure B1.**
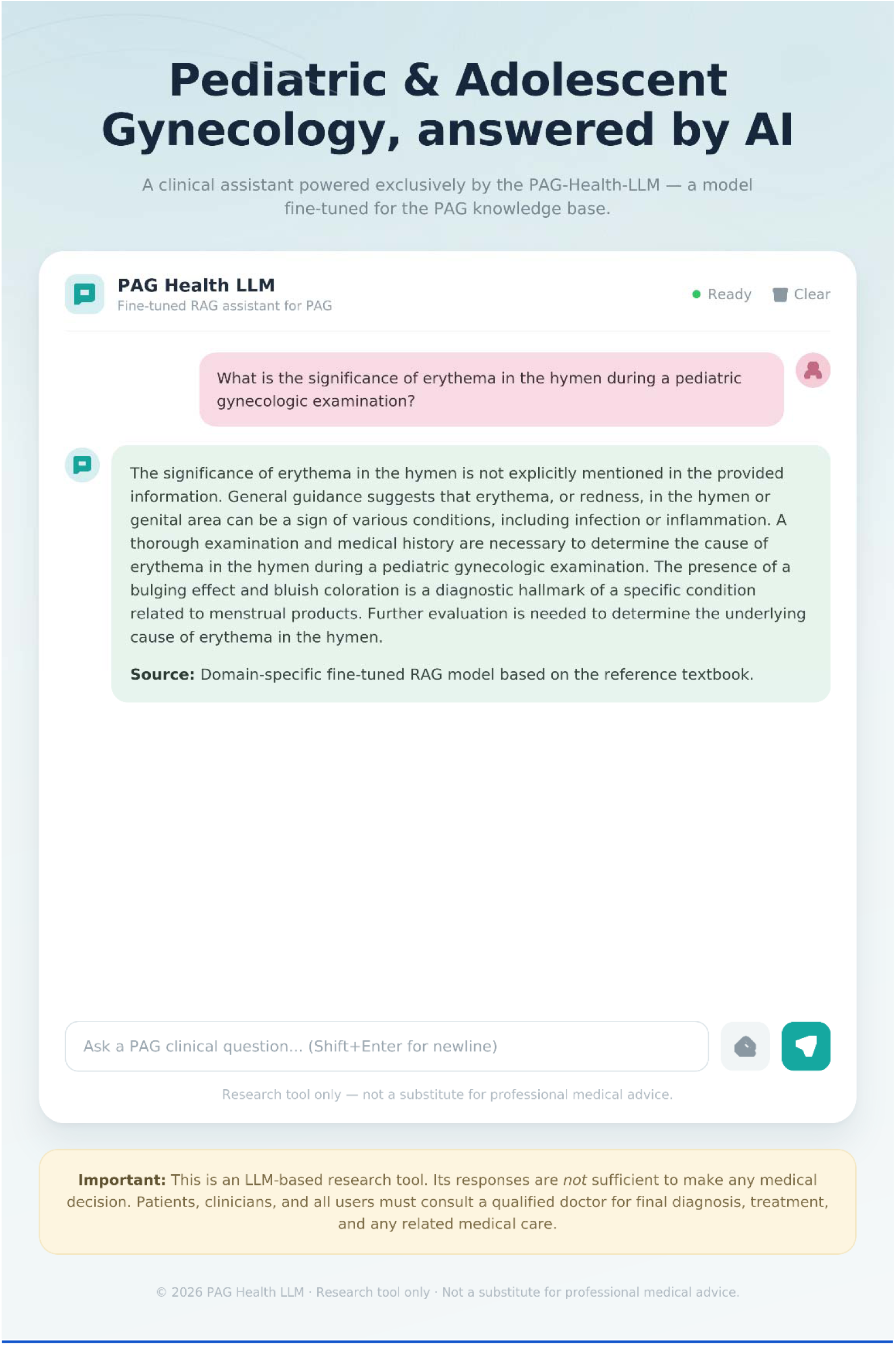
Live response from the deployed PAG-Health-LLM web application to test question 1: “What is the significance of erythema in the hymen during a pediatric gynecologic examination?” The model produces a concise citation-anchored response consistent with the textbook reference shown in Supplementary Table S1.

